# SARS-CoV-2 aerosol transmission in schools: the effectiveness of different interventions

**DOI:** 10.1101/2021.08.17.21262169

**Authors:** Jennifer Villers, Andre Henriques, Serafina Calarco, Markus Rognlien, Nicolas Mounet, James Devine, Gabriella Azzopardi, Philip Elson, Marco Andreini, Nicola Tarocco, Claudia Vassella, Olivia Keiser

**Author notes:** These authors contributed equally (Jennifer Villers, Andre Henriques).

## Abstract

**Background:** Indoor aerosol transmission of SARS-CoV-2 has been widely recognized, especially in schools where children remain in closed indoor spaces and largely unvaccinated. Measures such as strategic natural ventilation and high efficiency particulate air (HEPA) filtration remain poorly implemented and mask mandates are often progressively lifted as vaccination rollout is enhanced.

**Methods:** We adapted a previously developed aerosol transmission model to study the effect of interventions (natural ventilation, face masks, HEPA filtration, and their combinations) on the concentration of virus particles in a classroom of 160 m^3^ containing one infectious individual. The cumulative dose of viruses absorbed by exposed occupants was calculated.

**Results:** The most effective single intervention was natural ventilation through the full opening of six windows all day during the winter (14-fold decrease in cumulative dose), followed by the universal use of surgical face masks (8-fold decrease). In the spring/summer, natural ventilation was only effective (≥ 2-fold decrease) when windows were fully open all day. In the winter, partly opening two windows all day or fully opening six windows at the end of each class was effective as well (≥ 2-fold decrease). Opening windows during yard and lunch breaks only had minimal effect (≤ 1.2-fold decrease). One HEPA filter was as effective as two windows partly open all day during the winter (2.5-fold decrease) while two filters were more effective (4-fold decrease). Combined interventions (i.e., natural ventilation, masks, and HEPA filtration) were the most effective (≥ 30-fold decrease). Combined interventions remained highly effective in the presence of a super-spreader.

**Conclusions:** Natural ventilation, face masks, and HEPA filtration are effective interventions to reduce SARS-CoV-2 aerosol transmission. These measures should be combined and complemented by additional interventions (e.g., physical distancing, hygiene, testing, contact tracing, and vaccination) to maximize benefit.

## Introduction

Although children of all ages can be infected by and transmit SARS-CoV-2,^1^ it was initially thought that children did not play a major role in the transmission of the virus because they generally developed less severe symptoms and were more often asymptomatic than adults.^2^ However, many studies have described outbreaks in school settings,^3-8^ demonstrating that schools can contribute to the community spread of COVID-19. This has become more evident in England as the delta variant spread the fastest among secondary school-age children and young adults (17-24 years old), partly due to low vaccination rates and to the lack of mitigation measures in educational establishments.^9^

A major issue is the high risk of transmission of SARS-CoV-2 by airborne particles. This refers to the dispersion of the virus in small, invisible droplet nuclei that are generated when an infectious person exhales, talks, shouts, coughs, sneezes, or sings.^10^ These airborne particles, along with respiratory droplets, have the highest viral concentration in close proximity of an infected person although they can accumulate over time in poorly ventilated indoor spaces, floating in the air for minutes to hours. If another person inhales them, the virus can deposit on the surfaces of the respiratory tract and initiate infection of that person,^11^ making indoor spaces especially dangerous as infection can occur indirectly and over long distances.

Schools appear to be favorable places for SARS-CoV-2 transmission since children spend most of the day in a crowded and poorly ventilated space.^12^ Therefore, it is necessary to implement measures that can reduce the risk of aerosol transmission. Recommendations to keep schools safe vary widely from one region to another. In Europe, recommendations often focus on physical distancing and disinfection measures.^13-16^ When ventilation is mentioned, the focus is often on natural ventilation where the use of CO2 sensors is sometimes recommended to evaluate the level of ventilation.^17^ In contrast, the US CDC recommends the use of portable filtration devices in places where optimal natural ventilation cannot be reached.^18^ A similar divide exists regarding mask mandates where the US recommends all children and school personnel to wear a face mask, while most countries in Europe recommend it for older age groups only.^13, 15, 19^.

Regrettably, physical distancing and disinfection alone do not provide the required protection against long-range airborne transmission, so measures must also be taken to reduce airborne concentrations and overall potential viral dose absorbed by exposed individuals. Such measures include reducing crowding and time spent indoors, natural ventilation, the use of face masks, and the use of portable high-efficiency particulate air (HEPA) filtration systems.^10, 20^ This type of air filter can remove at least 99.97% of dust, pollen, mold, bacteria, viruses, and any airborne particles with a size of, at least, 0.3 microns (µm).

The objective of this paper is to assess the effectiveness of different interventions aimed at improving ventilation and reducing the risk of SARS-CoV-2 infection in a typical classroom setting using the COVID Airborne Risk Assessment (CARA) tool developed at the European Organization for Nuclear Research (CERN) by Henriques et al.,^21^ and to provide recommendations for decision makers that can improve schools’ safety.

## Methods

We used the COVID Airborne Risk Assessment (CARA) tool^21^ to assess the impact of different interventions on the concentration of virus particles (i.e., virions) in a classroom of 160 m^3^ containing one infectious occupant with COVID-19. CARA uses a physical model developed to simulate the concentration of virus particles in an enclosed indoor volume. Based on this, the cumulative dose of viruses absorbed by exposed occupants is calculated, which could be used to predict the probability of on-site transmission. The interventions modeled include different levels of natural ventilation, the universal use of surgical face masks, high efficiency particulate air (HEPA) filtration, as well as different combinations of these (see table 1). Each intervention is compared to a baseline scenario in which all windows are closed, no one is wearing a mask, and no filters have been installed.

**Table 1.**
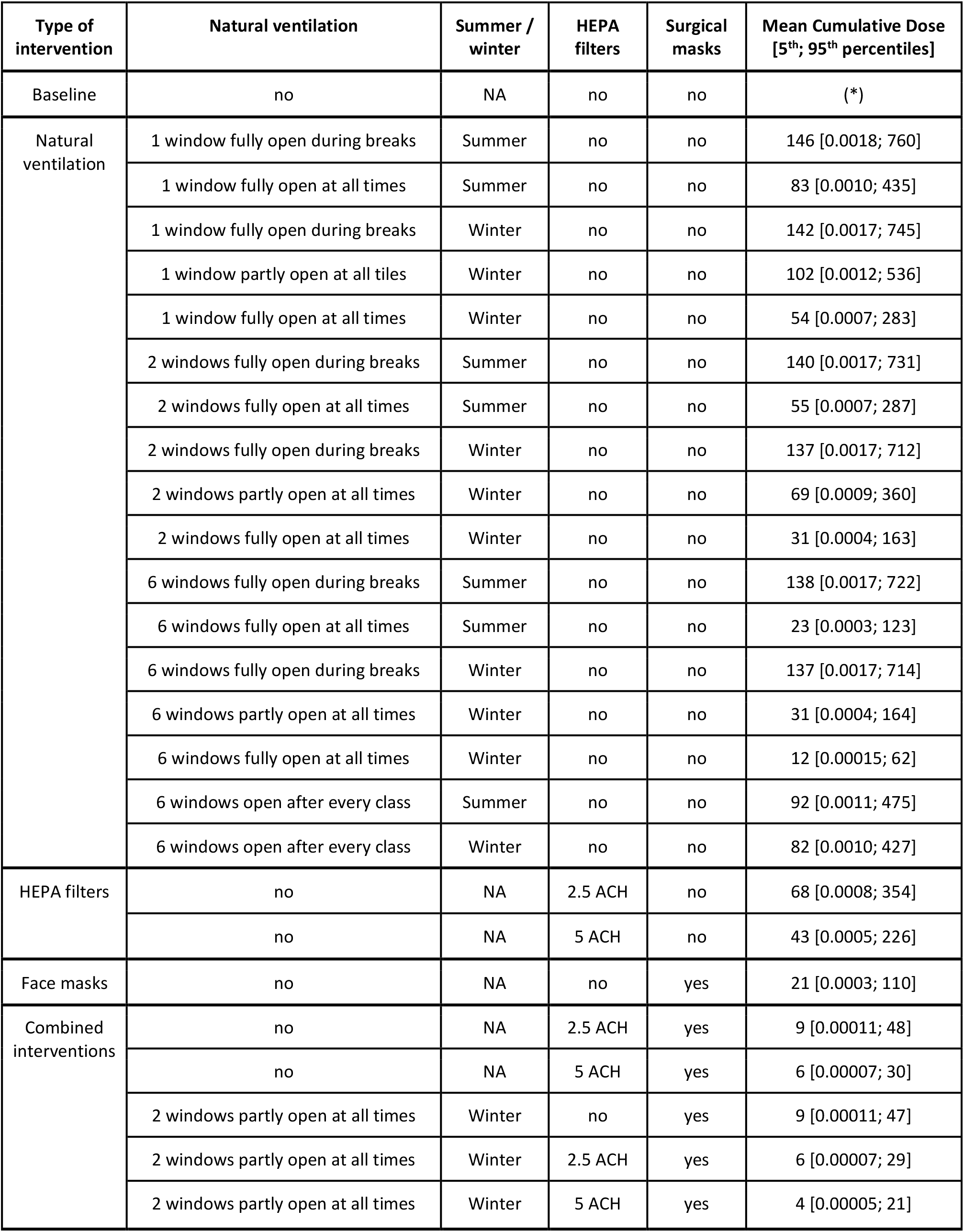
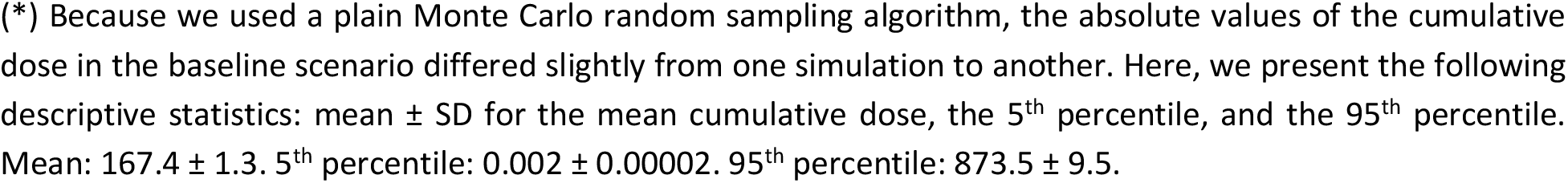
List of interventions.

| Type of intervention | Natural ventilation | Summer / winter | HEPA filters | Surgical masks | Mean Cumulative Dose [5<sup>th</sup>; 95<sup>th</sup> percentiles] |
| --- | --- | --- | --- | --- | --- |
| Baseline | no | NA | no | no | (*) |
| Natural ventilation | 1 window fully open during breaks | Summer | no | no | 146 [0.0018; 760] |
|  | 1 window fully open at all times | Summer | no | no | 83 [0.0010; 435] |
|  | 1 window fully open during breaks | Winter | no | no | 142 [0.0017; 745] |
|  | 1 window partly open at all times | Winter | no | no | 102 [0.0012; 536] |
|  | 1 window fully open at all times | Winter | no | no | 54 [0.0007; 283] |
|  | 2 windows fully open during breaks | Summer | no | no | 140 [0.0017; 731] |
|  | 2 windows fully open at all times | Summer | no | no | 55 [0.0007; 287] |
|  | 2 windows fully open during breaks | Winter | no | no | 137 [0.0017; 712] |
|  | 2 windows partly open at all times | Winter | no | no | 69 [0.0009; 360] |
|  | 2 windows fully open at all times | Winter | no | no | 31 [0.0004; 163] |
|  | 6 windows fully open during breaks | Summer | no | no | 138 [0.0017; 722] |
|  | 6 windows fully open at all times | Summer | no | no | 23 [0.0003; 123] |
|  | 6 windows fully open during breaks | Winter | no | no | 137 [0.0017; 714] |
|  | 6 windows partly open at all times | Winter | no | no | 31 [0.0004; 164] |
|  | 6 windows fully open at all times | Winter | no | no | 12 [0.00015; 62] |
|  | 6 windows open after every class | Summer | no | no | 92 [0.0011; 475] |
|  | 6 windows open after every class | Winter | no | no | 82 [0.0010; 427] |
| HEPA filters | no | NA | 2.5 ACH | no | 68 [0.0008; 354] |
|  | no | NA | 5 ACH | no | 43 [0.0005; 226] |
| Face masks | no | NA | no | yes | 21 [0.0003; 110] |
| Combined interventions | no | NA | 2.5 ACH | yes | 9 [0.00011; 48] |
|  | no | NA | 5 ACH | yes | 6 [0.00007; 30] |
|  | 2 windows partly open at all times | Winter | no | yes | 9 [0.00011; 47] |
|  | 2 windows partly open at all times | Winter | 2.5 ACH | yes | 6 [0.00007; 29] |
|  | 2 windows partly open at all times | Winter | 5 ACH | yes | 4 [0.00005; 21] |

(\*) Because we used a plain Monte Carlo random sampling algorithm, the absolute values of the cumulative dose in the baseline scenario differed slightly from one simulation to another. Here, we present the following descriptive statistics: mean ± SD for the mean cumulative dose, the 5<sup>th</sup> percentile, and the 95<sup>th</sup> percentile. Mean: $167.4 \pm 1.3$ . 5<sup>th</sup> percentile: $0.002 \pm 0.00002$ . 95<sup>th</sup> percentile: $873.5 \pm 9.5$ .

This study focuses on the ‘long-range’ airborne transmission route that assumes a well-mixed box model with a homogeneous viral concentration in the room. The model follows a probabilistic approach to deal with uncertainties of variables such as viral load of infected occupant(s) or breathing rate.^21^ For each intervention, 200,000 Monte Carlo simulations were performed. For each simulation, the viral load, breathing rate, and emission concentration were randomly sampled from distributions (see Fig S1).

### CARA has four main modules outlined below and described in detail elsewhere:^21^

1. The emission rate of viruses from the infected person’s mouth or nose (virions h^-1^, Figure S1a). This parameter is given by the volumetric count of respiratory fluid, in the form of aerosols, exhaled from the infected host per volume of breath (mL m^-3^) multiplied by its breathing rate (m^3^ h^-1^) and by the viral load (virions mL^-1^). For each Monte Carlo simulation, the viral load is randomly sampled from a distribution that has been determined by RT-PCR assays of nasopharyngeal swabs (Figure S1b),^22^ assuming an RNA-to-virus particle (virion) ratio of 1:1. The breathing rate is also randomly sampled from a distribution that depends on physical activity.^21^ Here, we assume that the infectious individual is standing, which could correspond to the teacher in the classroom (Figure S1c). The volumetric particle emission concentration is determined using the BLO model from Johnson et al.^23^ for different expiratory/vocal activities and includes the effect of masks. Here, we assume that the infectious individual is talking. The outward effectiveness of masks depends on the particle size, ranging from 35% for small particles to 80% for particles of ≥ 3 µm diameter (Figure S1d).^21^ From Figures S1a and S1b, we can already observe that the viral emission rate distribution is mostly influenced by the viral load distribution.
2. The dynamic concentration of viral particles in the air over the exposure time (virions m^-3^). Elementary specifications such as the volume of the room (160 m^3^) and its occupancy schedule are used as inputs. The concentration of viral particles will be largely governed by the viral removal rate which includes the effect of natural ventilation, air filtration, gravitational settlement of particles and the decay rate of aerosolized infectious viruses over time.^24, 25^ The effectiveness of natural ventilation is governed by the openable window area and by the difference between indoor and outdoor temperatures (discussed below). The effectiveness of air filtration depends on the number of filters and their effectiveness (discussed below). Other parameters are listed in table S1.
3. The cumulative absorbed dose of virus particles inhaled by an exposed host (virions). The dose is calculated by integrating the concentration profile over the total exposure time (in a stepwise function) and the effect of other parameters such as the physical activity of the occupants, the efficiency of masks of the exposed persons and an aerosol deposition factor in the respiratory tract.
4. The probability of on-site transmission, i.e., the probability that one susceptible exposed person gets infected, based on the absorbed dose. At the time of writing, the dose-response relationship for persons exposed to aerosolized SARS-CoV-2 viruses is not known to the authors. A few studies with other coronaviruses suggest an exponential response,^26^ meaning that a slight reduction in the inhaled dose would relate to an exponential reduction in the probability of contracting the disease, independently of the infective dose for SARS-COV-2. Preliminary experimental studies on SARS-CoV-2 suggest an infection dose between 10 - 1000 infectious virions.^27^ However, due to the high variability in infectious dose between SARS-CoV-2 variants, we only compared the relative effectiveness of different indoor preventive measures on the cumulative dose absorbed.

For natural ventilation, we considered the single-sided opening of one, two, or six windows (of 0.96 m^2^ each) all day long, during lunch and yard breaks, or for 10 minutes at the end of each class (in addition to breaks). Six windows correspond to an openable window area of 5.76 m^2^ (the average measured in Switzerland^28^). The opening width of the windows was set to 60 cm (fully open) or 20 cm (slightly open). School days were divided into 8 periods of 45 minutes with a 60-minute lunch break in the middle of the day and two 30-minute yard breaks (one in the morning and one in the afternoon). The occupants of the classroom were assumed to remain the same throughout the day and to leave the class during lunch and yard breaks. Since natural ventilation is influenced by the difference between indoor and outdoor temperatures, we simulated two different seasons: spring/summer—with an outdoor temperature of 18°C—and winter—with an outdoor temperature of 5°C. In both scenarios, the indoor temperature was set constant at 22°C.

For air filtration, we considered the use of one or two HEPA filtration devices, each delivering a flow rate up to 400 m^3^ h^-1^ of clean air (calculated based on a particle removal objective of at least 80% in 20 minutes that would yield an exchange rate of 2.5 air changes per hour, ACH). We assumed that the devices were strategically positioned in the room to ensure a homogenous filtering of the entire volume at occupant height (i.e., 1 to 1.8 m from the floor). In a typical square-like classroom, this would correspond to the center of the room.

For each intervention presented in table 1, we plotted the mean viral concentration and mean cumulative dose (Figures 1-3) as well as the full range of cumulative doses (Figure S2). The values for the mean cumulative doses as well as 5^th^ and 95^th^ percentiles are presented in table 1. We also generated heatmaps comparing the effectiveness of different interventions based on the relative amount by which they decrease the cumulative dose (Figure 1a-b).

**Figure 1.**
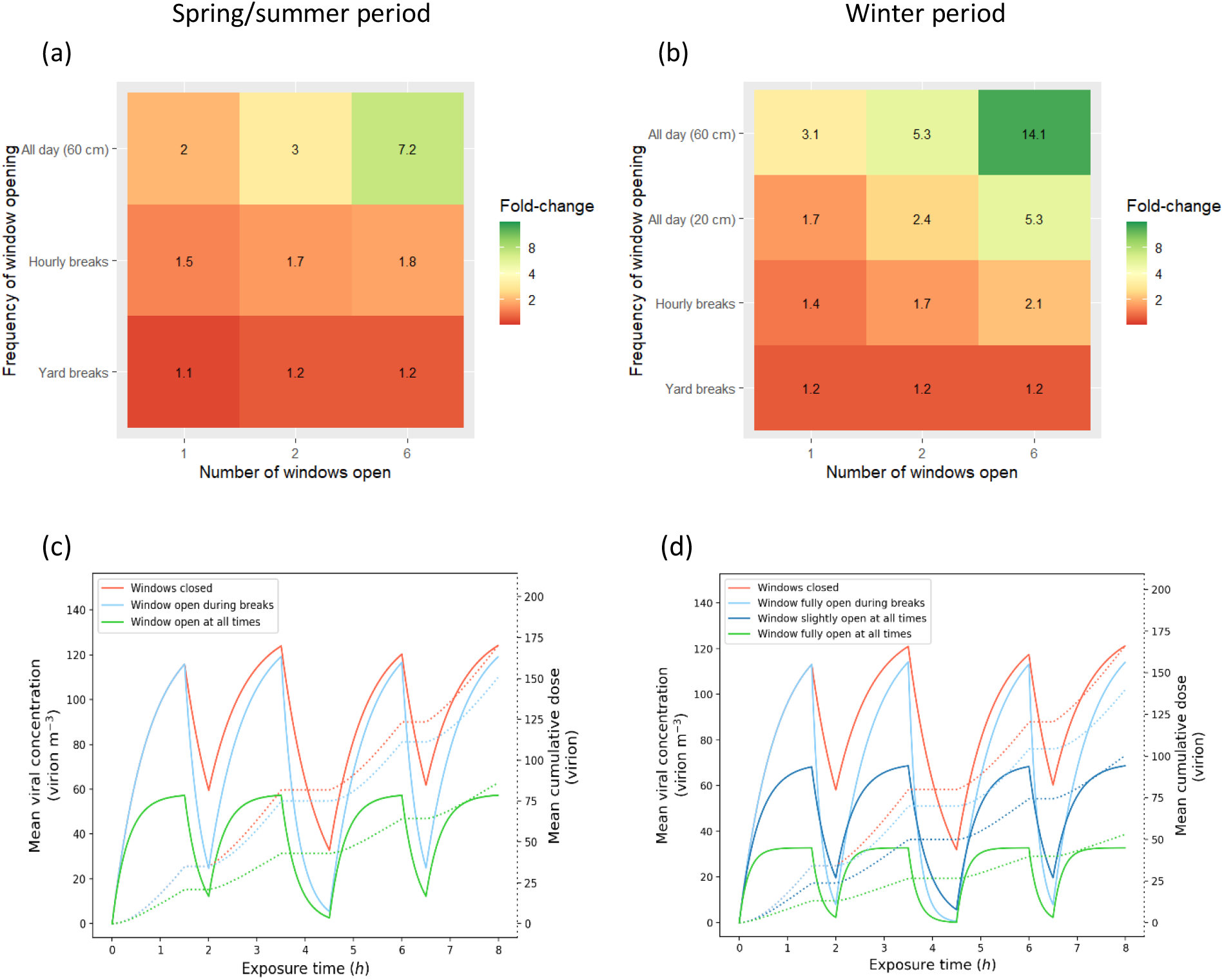
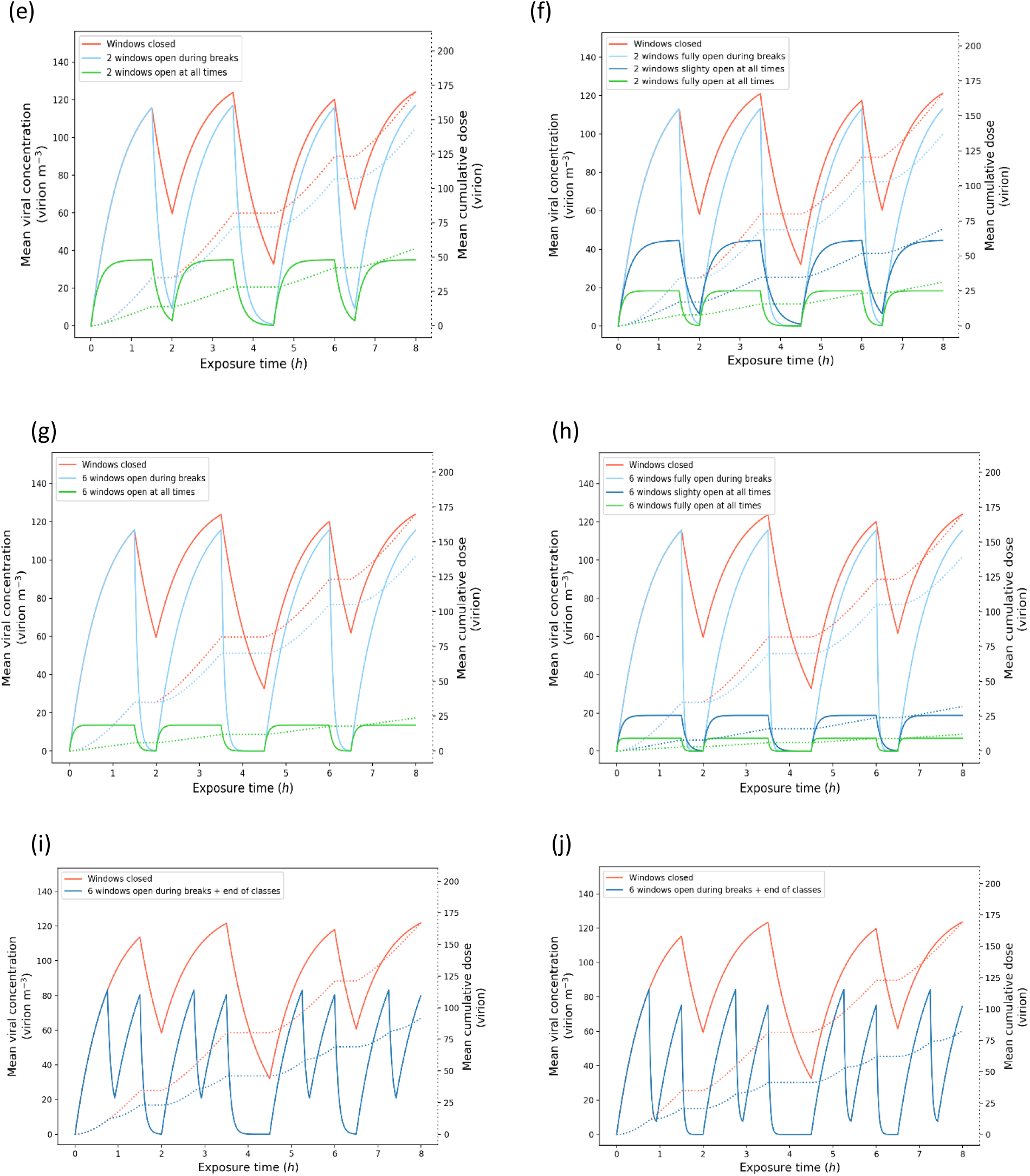
Natural ventilation. **(a-b)** Heatmaps of the relative effectiveness of natural ventilation strategies. Fold-decreases in cumulative dose absorbed (compared to the baseline scenario where windows are closed) are displayed as colors ranging from red to green as shown in the key. **(a)** Spring/summer simulations. **(b)** Winter simulations. **(c-j)** Estimated mean viral concentration profiles over the exposure time and consequent estimated mean cumulative dose of virions absorbed by the exposed hosts. The solid lines represent the concentration (left y-axis), and the dotted lines represent the cumulative dose (right y-axis). The red lines show the results for the baseline scenario, the blue and green lines show the results for different natural ventilation scenarios. **(c-h)** Light blue: windows are fully open during breaks only. Green: windows are fully open (60 cm) all day. Dark blue: windows are partly open (20 cm) all day. **(c-d)** Results with one window. **(c)** Spring/summer simulations. **(d)** Winter simulations. **(e-f)** Results with two windows. **(e)** Spring/summer simulations. **(f)** Winter simulations. **(g-h)** Results with 6 windows **(g)** Spring/summer simulations. **(h)** Winter simulations. **(i-j)** Results with 6 windows opened at the end of each class. **(i)** Spring/summer simulations. **(j)** Winter simulations.

## Results

When looking at the full range of simulations for the cumulative dose absorbed by a susceptible person (Figure S2), we observed that it spans several orders of magnitude (e.g., from 0.0005 to over 2,500 virions in the baseline scenario). This outcome was not surprising considering the wide distribution of viral loads in the infected host population (ranging from 100 to 10 billion virions mL^-1^, Figure S1b). When using plain Monte Carlo random sampling algorithm, the absolute values of the cumulative dose can differ slightly from one simulation to another. However, the effectiveness of the interventions can be determined by studying the relative difference of their respective cumulative dose results, which is conserved throughout the simulations (Figure S2 and Table 1).

The effectiveness of natural ventilation was dependent on the number of windows open, the duration and frequency of these openings, and the difference between indoor and outdoor temperature (Figure 1). Opening windows during yard and lunch breaks only had minimal effect on the cumulative dose of virions absorbed, with decreases in cumulative dose ranging from 1.1-fold to 1.2-fold depending on the season and number of windows open. In contrast, opening one, two, or six windows all day long during spring/summer decreased the cumulative dose absorbed 2-fold, 3-fold, and 7-fold, respectively, compared to the baseline scenario. Keeping windows open all day was most effective in the winter, with a 3-fold decrease in the cumulative dose absorbed when one window was fully open, a 5-fold decrease when two windows were fully open, and a 14-fold decrease when six windows were fully open. Since leaving windows wide open during the heating season is unacceptable (waste of heating energy), we tested a 20 cm opening all day long leading to a 1.7-fold decrease in the cumulative dose when one window was open, a 2.4-fold decrease when two windows were open, and a 5-fold decrease when six windows were open. We also tested the opening of six windows at the end of each class leading to a 1.8-(summer) and 2.1-fold (winter) decrease in cumulative dose.

The second intervention simulated was the use of HEPA filtration devices. We tested the recommended 5 air changes per hour (ACH),^29^ where two filters per classroom are needed (HEPA device each delivering a flow rate up to 400 m^3^h^-1^ of clean air), and an intermediate 2.5 ACH corresponding to one filter (Figure 2a). The 2.5 ACH option was as effective as two windows 20-cm open all day long during winter (2.5-fold decrease). The 5 ACH option was even more effective with a 4-fold decrease in the cumulative dose absorbed. The universal use of face masks was twice as effective, with an 8-fold decrease in the cumulative dose absorbed (Figure 2b).

**Figure 2.**
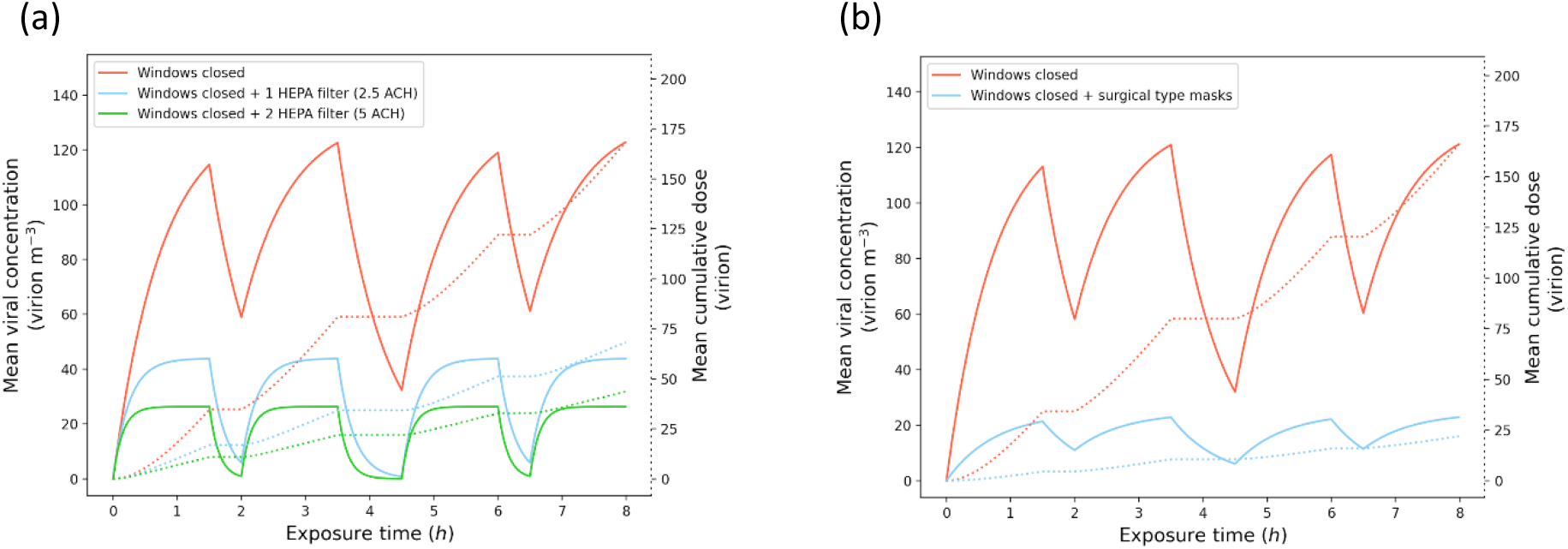
(a) HEPA filtration and (b) surgical face masks. Estimated mean viral concentration profiles over the exposure time and consequent estimated mean cumulative dose of virions absorbed by the exposed hosts. The solid lines represent the concentration (left y-axis), and the dotted lines represent the cumulative dose (right y-axis). The red lines show the results for the baseline scenario with closed windows and no filter. **(a)** The blue and green lines show the results for HEPA filtration scenarios with 2.5 and 5 air changes per hour (ACH), respectively. **(b)** The blue lines show the results for a scenario without natural ventilation nor HEPA filters where both the infectious and exposed individuals wear surgical face masks.

Combined interventions were generally the most effective. For instance, adding surgical face masks to natural ventilation decreased the cumulative dose absorbed 8-fold compared to ventilation alone, reducing the total cumulative dose 19-fold compared to the baseline scenario (Figure 3a-b). Similarly, combining surgical facemasks and HEPA filtration led to an 18-fold decrease for 2.5 ACH and to a 29-fold decrease for 5 ACH. Combining natural ventilation, surgical face masks and HEPA filtration reduced the cumulative dose 30-fold for 2.5 ACH and 40-fold for 5 ACH compared to the baseline scenario (Figure 3b).

**Figure 3.**
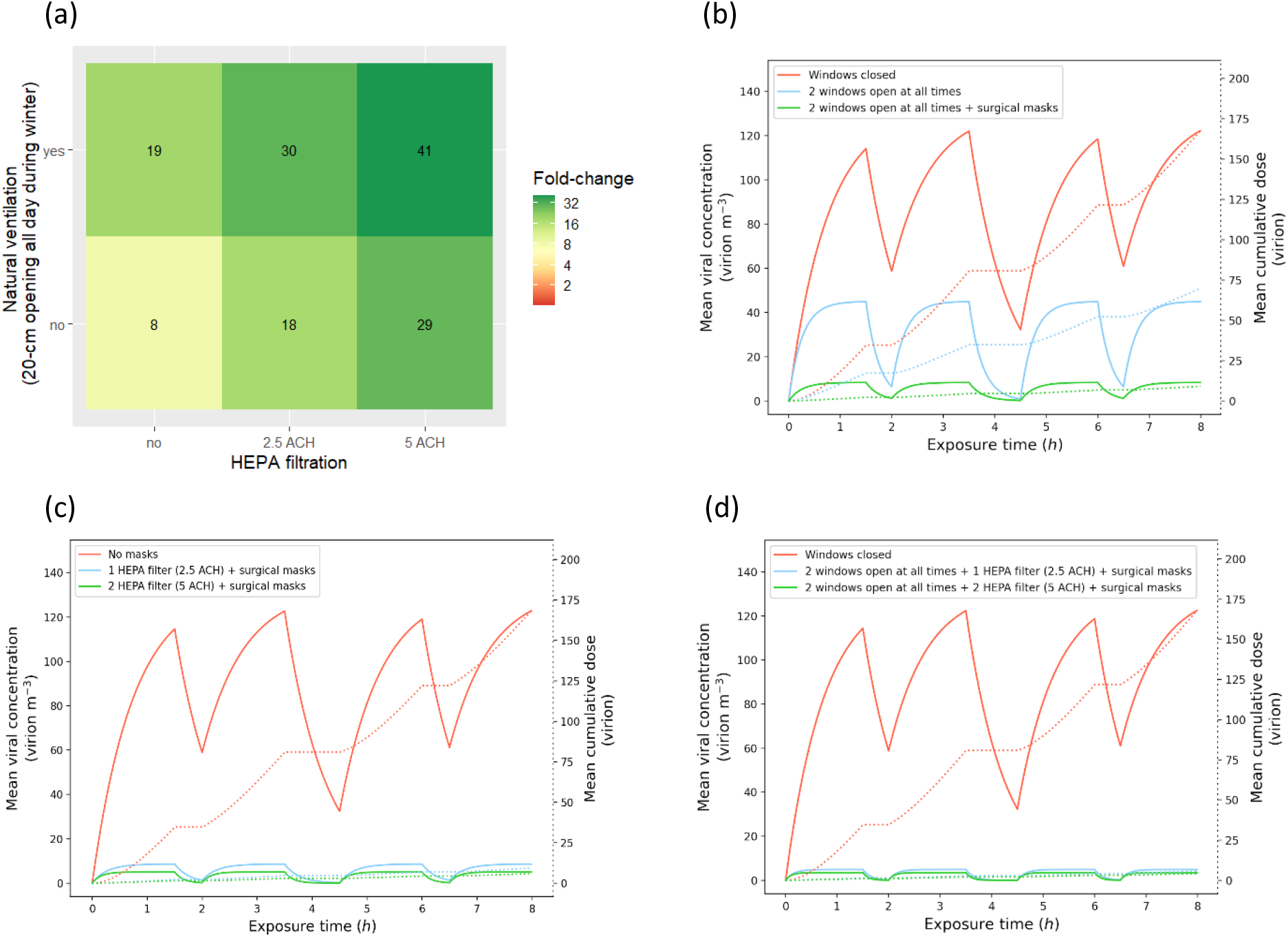
Combinations of interventions. **(a)** Heatmap of the relative effectiveness of combined interventions. Fold-decreases in cumulative dose absorbed (compared to the baseline scenario where windows are closed) are displayed as colors ranging from red to green as shown in the key. **(b-d)** Estimated mean viral concentration profiles over the exposure time and consequent estimated mean cumulative dose of virions absorbed by the exposed hosts, for different scenarios. The solid lines represent the concentration (left y-axis), and the dotted lines represent the cumulative dose (right y-axis). The red lines show the results for the baseline scenario with closed windows, no HEPA filtration, and no face masks, the blue and green lines show the results for different combinations of interventions. **(b)** Results with natural ventilation (two windows 20-cm open all day long during winter) without surgical face masks (blue) and with surgical face masks (green). **(b)** Results with face masks and HEPA filtration (blue: 2.5 ACH; green: 5 ACH). **(c)** Results with natural ventilation (two windows 20-cm open all day long during winter) and face masks and HEPA filtration (blue: 2.5 ACH; green: 5 ACH).

Since around 15% of individuals are responsible for 80% of secondary transmissions,^30^ we evaluated the impact of interventions when the infectious individual is a super-spreader. Since the cumulative dose absorbed by an exposed occupant in our model is strongly influenced by the viral load of the infectious occupant, we looked at the highest percentiles of the cumulative dose distributions (Figure S2). Under baseline conditions, the 95^th^ percentile cumulative dose was 5-fold higher than the mean (∼900 virions) and the 99^th^ percentile 16-fold higher (∼2,700 virions). When natural ventilation was combined with the universal use of surgical face masks (Figure S2k), the 99^th^ percentile cumulative dose remained lower than the mean cumulative dose under baseline conditions (144 virions). When all three interventions were combined (Figure S2m), the 99^th^ percentile cumulative dose remained below 100 virions (91 virions for 2.5 ACH and 67 virions for 5 ACH). These results suggest that combined interventions remain highly effective against super-spreaders.

## Discussion

Using the COVID Airborne Risk Assessment (CARA) modeling tool,^21^ we found that surgical face masks, HEPA filters, and strategic natural ventilation are effective strategies to reduce the cumulative dose of virions absorbed by exposed individuals in a classroom setting. We also showed that these interventions have cumulative effects and should be implemented together for maximal benefit. This is in agreement with the results of a recent study of the US CDC that showed—using a breathing aerosol source simulator—that HEPA filtration combined with face masks can reduce the mean aerosol concentration by 90%.^31^

Opening windows only during yard and lunch breaks was the least effective intervention and should not be recommended. We found natural ventilation to be most effective in the winter. This result was expected since the fresh air flow for single-sided natural ventilation is proportional to 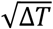, with ∆*T* representing the difference between outdoor and indoor temperature.^21^ As a consequence, in the spring/summer (assuming ∆*T* = 4°C), natural ventilation was only effective when windows were fully open all day long. Conversely, less extreme ventilation strategies such as a 20 cm opening of two windows all day or the full opening of six windows at the end of each class reduced by at least half the cumulative dose absorbed in winter (assuming ∆*T* = 17°C).

While natural ventilation through the opening of windows at all times is an effective strategy to decrease the concentration of virions in the air, it can be inadvisable when outdoor temperatures are too extreme, when the outdoor air is too polluted, or when there is too much noise. In addition, when the outdoor temperature equals the indoor temperature, natural ventilation is less effective. This prompted us to consider alternative strategies to reduce virion concentration such as the use of high efficiency particulate air (HEPA) filtration devices, which performance is unaffected by the season. The effectiveness of HEPA filters is measured in air changes per hour (ACH) of the device’s clean air delivery rate^32^ and must be calculated based on the volume of the room and the specifications of the device. However, “HEPA” *per se* is not a certification label and might not be enough to guarantee the desired performance. We suggest to install filters labeled H13 (≥99.95% efficiency) or H14 (≥99.995% efficiency), according to EN 1822 standard.^33^

We found HEPA filtration to be an effective strategy to decrease the cumulative dose of virions absorbed by exposed individuals, with the 2.5 ACH option leading to a 2.5-fold decrease in cumulative dose and the 5 ACH option to a 4-fold decrease. The main disadvantage of HEPA filters compared with face masks or natural ventilation strategies is their cost—H13 and H14 HEPA filters can be expected to cost USD ∼1000 and filters need to be replaced at least every two years, or according to manufacturer’s instruction, at a cost of USD ∼300.

CO_2_ sensors, which are more affordable (USD ∼160), can be used to assess the level of natural ventilation in view of identifying rooms which are poorly ventilated and would most benefit from the addition of HEPA filters, favoring a targeted investment for the school structure. However, there is no direct correlation between CO_2_ levels and virion concentration since the latter is influenced by the number of infected hosts and their viral load. While no CO_2_ concentration can guarantee occupants’ safety, current recommendations are to keep CO_2_ levels below 1000 ppm to reduce the risk of transmission of SARS-CoV-2 during sedentary activities such as classes.^17, 34, 35^ Rooms where that threshold cannot be reached by natural ventilation should be prioritized for complementary measures such as HEPA filtration or the installation of a mechanical ventilation system. Levels as high as 4400 ppm have been observed in densely occupied, poorly ventilated spaces such as schools, highlighting the importance of establishing a ventilation strategy in classrooms.^10, 28^ Importantly, while HEPA devices filter particulate matter, they do not filter gaseous molecules. As a consequence, CO_2_ sensors cannot be used to assess the efficacy of HEPA filters and HEPA filtration devices should be combined with natural or mechanical ventilation to ensure that CO_2_ concentrations do not exceed levels that have been found to interfere with students’ performance. While CO_2_ levels are influenced by the number of people occupying the room, the viral concentration is only influenced by the number of infectious individuals, their viral load, and their physical and expiratory activities. In this paper, we assumed a homogenous viral concentration in a classroom containing one infectious individual, meaning that all other room occupants have an equal exposure to virions.

The universal use of surgical face masks was one of the most effective interventions to reduce the cumulative dose absorbed by the exposed individuals, second only to the full opening of six windows all day long during the winter. However, similar to HEPA devices, what is crucial is to guarantee the desired filtration performance with the proper certification. We suggest to use well fit surgical masks labeled with a material filtration performance of, at least, Type I (≥95% bacterial filtration efficiency), according to EN 14683 standard.^36^

Combined interventions—i.e., natural ventilation in combination with surgical face masks and HEPA filters—remained highly effective in the presence of a super-spreader. Heterogeneity in transmission is a characteristic feature of SARS-CoV-2, with around 15% of infected individuals responsible for 80% of secondary infections.^30^ This high heterogeneity in transmission has been traced back to heterogeneity in both the number of contacts and in individuals’ viral loads—with 2% of individuals harboring up to 90% of virions.^37^ This effect has been proposed as an explanation for the apparent discrepancies in the literature regarding transmission of SARS-CoV-2 in school settings—with most children not transmitting the disease to anyone and a handful being responsible for large outbreaks.^38^ In addition, viral load distributions were found to be similar between children, youths, and adults as well as between symptomatic and asymptomatic individuals^37^—with around half of transmissions occurring during the presymptomatic phase.^30^ Since highly contagious individuals cannot be identified based on age or symptoms, mitigation strategies such as ventilation and face masks are especially important in crowded, closed settings such as classrooms.

One important limitation of our model comes from the lack of consensus regarding the infectious dose. Hence, we cannot predict, on a quantitative level, what measures are sufficient to keep the occupants of the room safe. Nonetheless, reducing the number of inhaled virions will result in an exponential reduction in the probability of contracting the disease. Therefore, we can estimate the mean cumulative dose absorbed in each scenario and by how much it varies based on the interventions implemented. Furthermore, the infectious dose is hypothesized to vary depending on the SARS-CoV-2 variant. Since no significant change in viral load has been observed between the original variant and the alpha variant,^39^ the most likely hypothesis to explain the increased infectivity of the alpha variant is a genetic mutation in the spike protein enabling it to more effectively bind to the ACE2 receptor,^40^ thereby decreasing the amount of viral particles needed to infect a susceptible host. With the rise of the delta variant, which became dominant in England within months^8^ and is rapidly spreading across many countries, we can expect the infectious dose to decrease, making new outbreaks more difficult to contain. For example, in Bolton, UK, where the delta variant became first dominant, data suggested that the infection first spread among school-age children.^41^ The delta variant has already been identified in 195 outbreaks or clusters in primary and secondary schools in England from April 26 to June 13, 2021.^8^ Secondary school-age children had the highest overall infection rates in England during the first week of June before being overtaken by young adults (17-24 years old).^9^ Until vaccination reaches all age groups, mitigation strategies such as ventilation and face masks are especially warranted in unvaccinated and crowded populations in order to prevent the rise of new and potentially more dangerous SARS-CoV-2 variants.

Another major limitation of our model is the assumption of a homogenous viral concentration throughout the room, as it does not account for the proximity to the infectious individual. A well-mixed model has the advantage of simplicity and requires lower computing power, which can be an asset when performing a rapid assessment for each room in a school. However, it only evaluates the effectiveness of interventions against long-range) airborne transmission. Additional hygiene and distancing measures remain necessary to prevent short-range transmission. Among the interventions that we tested, the universal use of face masks was one of the most effective at preventing long-range transmission and is the only one that also protects against short-range transmission. Recently, several countries have lifted requirements for face coverings in secondary schools despite increasing infection rates in educational settings.^42, 43^ In an opinion piece, Gurdasani and colleagues “argued that this was ill-advised given the clear evidence for the role of children and schools in transmission of SARS-CoV-2 and the rise of the new (delta) variant”.^44^ Our results further support this position since universal face masks decreased 8-fold the cumulative dose absorbed by exposed individuals.

Besides primary and secondary education settings, the CARA tool can also be used to assess the effectiveness of mitigation measures in other settings, such as universities, other higher education institutions, and other indoor spaces. Beyond the current COVID-19 crisis, maintaining a proper level of ventilation in classrooms is also recommended for the children’s health and performance.^45^ Some of the most pathogenic viruses such as influenza, respiratory syncytial virus, adenovirus, coronavirus and measles are transmitted through aerosols, while elevated levels of CO_2_ have been found to interfere with intellectual concentration and, thus, students’ performance. Besides proper ventilation, HEPA filtration devices would also be useful beyond the pandemic as they might capture larger airborne particles such as pollen or outdoor atmospheric pollutants, which were found to be associated with increased susceptibility to respiratory diseases such as COVID-19.^46, 47^

## Data Availability

Access to the tool and model code can be obtained from the corresponding author.

https://gitlab.cern.ch/cara/cara

## Acknowledgements

The authors thank Sabina Rodriguez Velásquez for improving the use of English in the manuscript.

## Contributors

JV, AH, SC, and OK conceived and planned the numerical experiments. AH, MR, NM, JD, GA, PE, MA, and NT designed the model and the computational framework. AH carried out the simulations. JV, AH, SC, CV, and OK interpreted the results. JV and AH designed the figures. JV drafted the manuscript and AH, CV, and OK revised it critically. OK supervised the project. All authors reviewed and approved the final manuscript.

## Ethical approval

This research did not involve human or animal subjects, and ethical approval was therefore not required.

## Data sharing statement

CARA is an Open Source software under an APACHE License, Version 2.0. The code repository can be accessible here: https://gitlab.cern.ch/cara/cara. Other information can be obtained from the corresponding author.

