## Supplementary material for "SARS-CoV-2 aerosol transmission in schools: the effectiveness of different interventions"

### SUPPLEMENTARY INFORMATION

Table S1. Parameters for the concentration model

| Parameter | Value | Unit |
|---|---|---|
| Inactivation rate due to aerosol gravitational settlement | 0.45 | $h^{-1}$ |
| Inactivation rate due to inactivity decay in air | 0.63 | $h^{-1}$ |
| Aerosol deposition factor | 0.6 | - |
| Outward mask efficiency | [35-80] (*) | % |
| Inward mask efficiency | 50 | % |

(*) As a function of particle diameter

**Figure S1. Input parameters and associated result of the emission rate.** (a) Results of the Monte Carlo simulations for the determination of the emission rate distribution for an infected host talking, while undertaking light physical activity (i.e., standing). The vertical axis corresponds to the estimation of distribution Probability Density Function (PDF). The median emission rate value is 862 virion $h^{-1}$ with a mean (SD) of 2.7 (1.7) $\log_{10}$ (virion $h^{-1}$). The super-spreader range is represented by the values one standard deviation from the mean, where the emission rate increases to >25 100 virion $h^{-1}$. Results computed from a sample size of 200,000. (b) Viral load distribution resulting from 20,000 RT-PCR assays in the population.[22] (c) Distribution of the breathing rate while undertaking light physical activity. The same breathing rate distribution was used for the exposed children, shifted to a mean of 0.51m3/h to account for their level of physical activity while seated. (d) Size distribution particle emission while the host is talking, with and without surgical face mask.

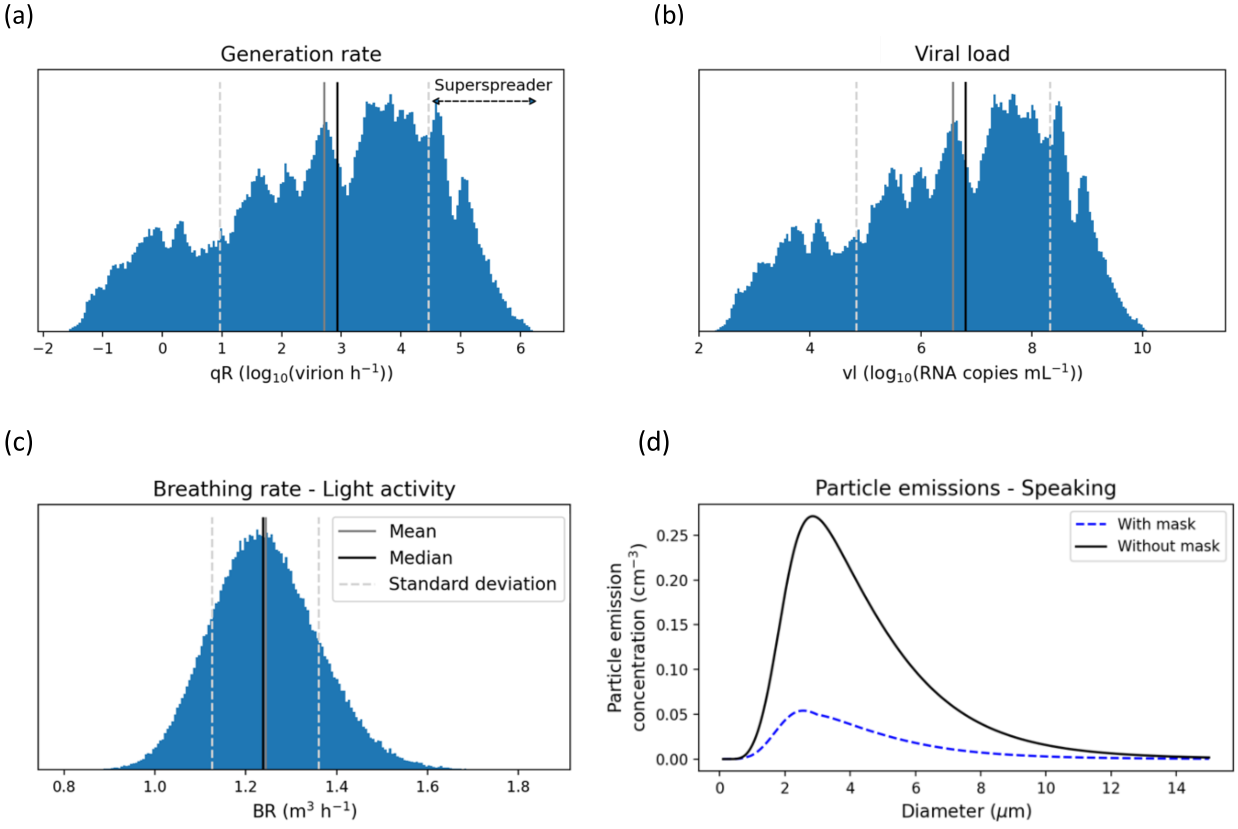

**Figure S2. Cumulative dose distributions.** Results from the 200,000 Monte Carlo simulations for each intervention.

(a) Natural ventilation spring/summer 1 window

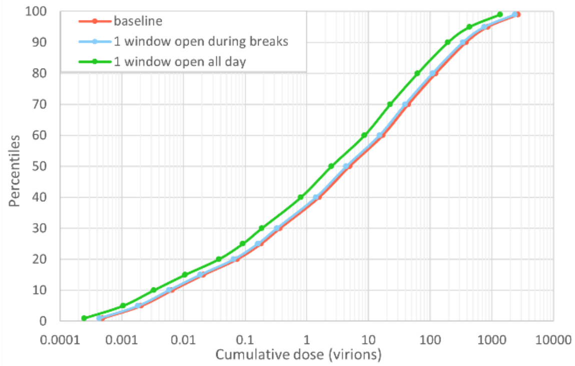


(b) Natural ventilation winter 1 window

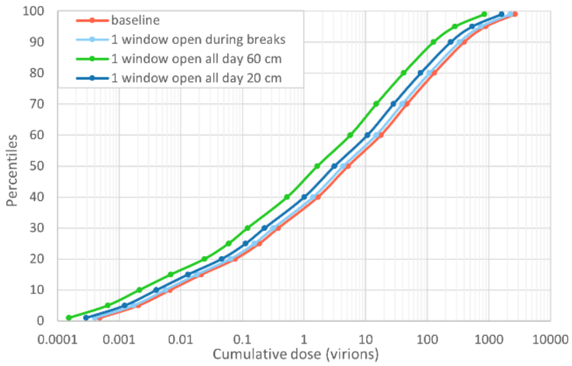


(c) Natural ventilation spring/summer 2 windows

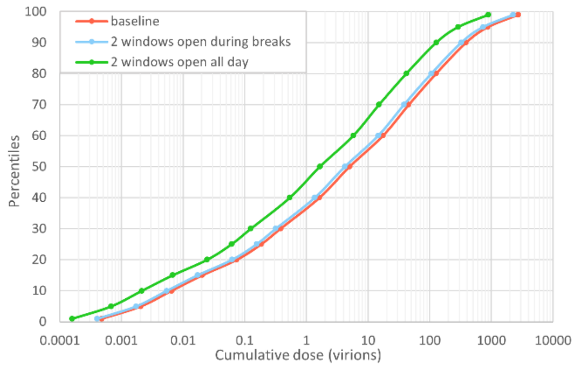


(d) Natural ventilation winter 2 windows

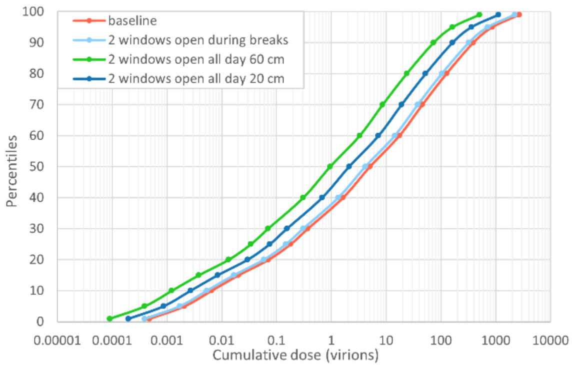


(e) Natural ventilation spring/summer 6 windows

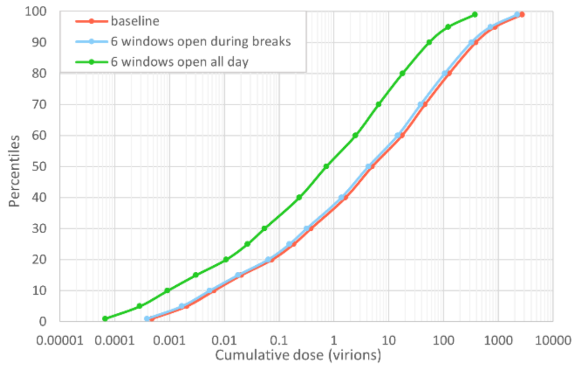


(f) Natural ventilation winter 6 windows

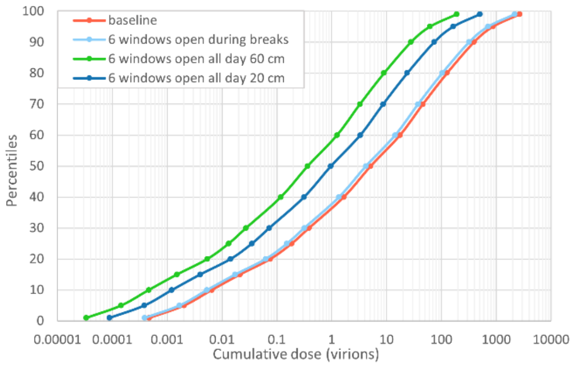

(g) Natural ventilation spring/summer every 45 min

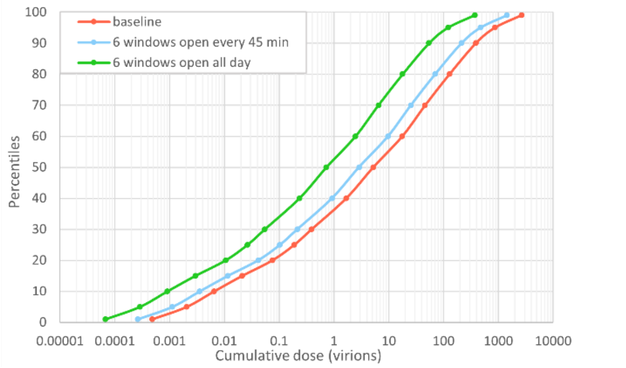


(h) Natural ventilation winter every 45 min

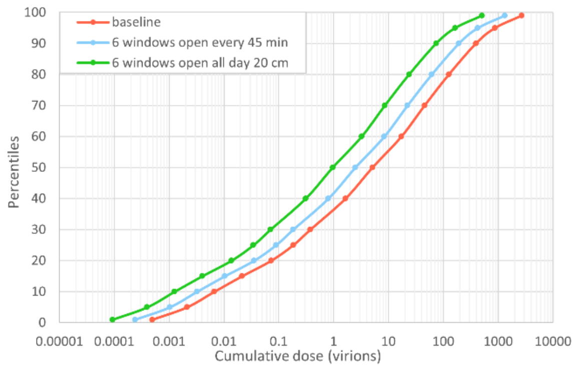


(i) HEPA filter

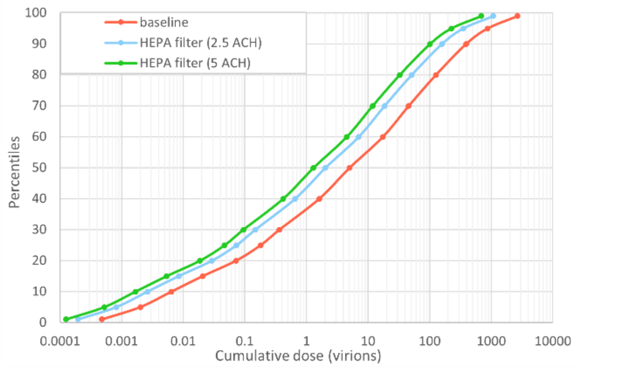


(j) Surgical face masks

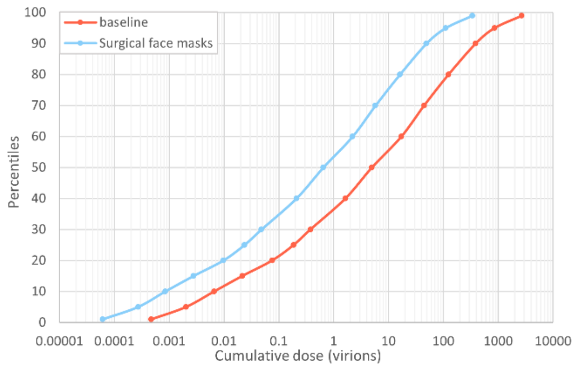


(k) Combined interventions

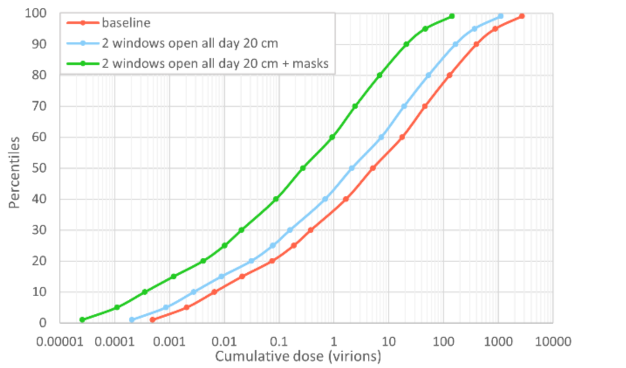


(l) Combined interventions

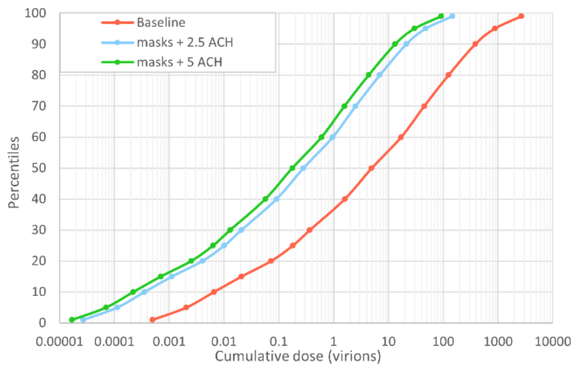


(m) Combined interventions

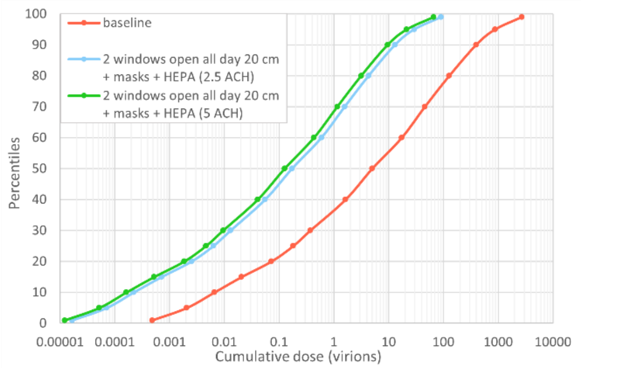
